# The physio-affective phenome of major depression is strongly associated with biomarkers of astroglial and neuronal projection toxicity which in turn are associated with peripheral inflammation, insulin resistance and lowered calcium

**DOI:** 10.1101/2022.07.04.22277246

**Authors:** Hussein Kadhem Al-Hakeim, Tabarek Hadi Al-Naqeeb, Abbas F. Almulla, Michael Maes

## Abstract

**Background:** Major depressive disorder (MDD) is characterized by elevated activity of peripheral neuro-immune and neuro-oxidative pathways, which may cause neuro-affective toxicity by disrupting neuronal circuits in the brain. No study has explored peripheral indicators of neuroaxis damage in MDD in relation to serum inflammatory and insulin resistance (IR) biomarkers, calcium, and the physio-affective phenome consisting of depressive, anxious, chronic fatigue, and physiosomatic symptoms.

**Methods:** Serum levels of phosphorylated tau protein 217 (P-tau217), platelet-derived growth factor receptor beta (PDGFR), neurofilament light chain (NF-L), glial fibrillary acidic protein (GFAP), C-reactive protein (CRP), calcium and the HOMA2-insulin resistance (IR) index were measured in 94 MDD patients and 47 controls.

**Results:** 61.1% of the variance in the physio-affective phenome (conceptualized as a factor extracted from depression, anxiety, fatigue and physiosomatic symptoms) is explained by the regression on GFAP, NF-L, P-tau2017, PDGFRβ and HOMA2-IR (all positively associated), and decreased calcium. In addition, CRP and HOMA2-IR predicted 28.9% of the variance in the neuroaxis index. We observed significant indirect effects of CRP and calcium on the physio-affective phenome which were partly mediated by the four neuroaxis biomarkers. Annotation and enrichment analysis revealed that the enlarged GFAP, P-tau217, PDGFR, and NF-L network was enriched in glial cell and neuronal projections, the cytoskeleton and axonal transport, including a mitochondrion.

**Conclusions:** Peripheral inflammation and IR may damage the astroglial and neuronal projections thereby interfering with mitochondrial transport. This toxicity, combined with inflammation, IR and lowered calcium, may, at least in part, induce the phenome of MDD.

## Introduction

Major depressive disorder (MDD) is the most prevalent mental disease in the world, with high morbidity and increased mortality and suicide risk (Ferrari et al., 2013; World Health Organization, 2016). With a lifetime prevalence of 10.8 percent (Lim et al., 2018), MDD is a widespread mental condition that affects over 300 million individuals all over the globe (Patel et al., 2016).

There is now evidence that MDD is characterized by activation of the immune-inflammatory response system (IRS) with activation of M1 macrophage, T helper (Th)-1 and Th-17 phenotypes and increased production of growth factors, including platelet derived growth factor (PDGF) (Maes and Carvalho, 2018; Maes et al., 2022a). Some cytokines and chemokines that are overproduced in MDD, such as interleukin (IL)-15, IL-17, IL-1β, IL-6, tumor necrosis factor (TNF)-α, interferon (IFN)-γ, CXCL8, CXCL10, and CCL5, have neurotoxic effects and may jeopardize neuronal functions, such as neurogenesis, neuroplasticity, receptor expression, axonogenesis, and dendritic sprouting (Maes et al., 2022a; Maes et al., 2009). Moreover, elevated levels of IL-6, IL-1β, and TNF-α induce an acute phase response in the liver, leading to increased production of C-reactive protein (CRP) (Sluzewska et al., 1996), which has additional toxic effects on endothelial cells, thereby increasing the permeability of the blood-brain barrier (BBB) and exacerbating the development of neurodegenerative and cerebrovascular processes (Belin et al., 2020; Ge et al., 2013; Hsuchou et al., 2012; Song et al., 2009; Windgassen et al., 2011).

Moreover, in MDD, the neuroimmunotoxic effects of these cytokines/chemokines and CRP may exacerbate the neurotoxic consequences of increased lipid and protein oxidation, aldehyde formation, and hypernitrosylation (Maes et al., 2022a). Diminished antioxidant and neurotrophic defenses, including decreased levels of brain-derived neurotrophic factor (BDNF), and a relative deficiency in the compensatory immunoregulaty system (CIRS) further contribute to enhanced neurotoxic effects of the IRS (Maes and Carvalho, 2018; Maes et al., 2022a; Maes et al., 2009; Mehterov et al., 2022). New precision nomothetic models of MDD demonstrate that IRS activation and subsequent affective neurotoxicity dictate not only the phenome of MDD, but also the recurrence of illness (ROI) and suicidal behaviors (Maes, 2022; Maes et al., 2021; Vasupanrajit et al., 2021).

Moreover, the IRS response in MDD is further enhanced by increased production of growth factors including PDGF which regulate chemotaxis, cell division, cell proliferation, and mitogen-activated protein kinase (MAPK) signaling pathways (3 items (human) - STRING interaction network (string-db.org) and, therefore, contribute to enhanced neurotoxicity in MDD (Maes et al., 2022a). The binding of PDGF to the PDGF receptor (PDGFR) induces downstream signaling leading not only to cell proliferation, migration, and differentiation (Andrae et al., 2008; Nicolas et al., 2013) but also to dysfunctions in synaptogenesis, neurogenesis, ligand-gated ion channels, and neurovascular and neurodegenerative processes (Sil et al., 2018). CSF levels of PDGFR, which is a pericyte biomarker (Chen et al., 2017), are elevated in dementia patients (Sweeney et al., 2020) and are associated with increased BBB permeability in moderate cognitive impairment patients (Nation et al., 2019).

On the basis of the IRS-affective neurotoxicity theory of MDD (Maes, 2008; Maes et al., 2022a; Schiepers et al., 2005a), it is essential to investigate whether biomarkers of neurotoxicity, damage to the neuroaxis, and brain injuries are involved in the pathophysiology of MDD and are associated with the IRS response. MDD and bipolar patients, for example, have considerably elevated plasma neurofilament light chain (NF-L) levels (Chen et al., 2022; Jakobsson et al., 2014), and treatment-resistant depression is associated with elevated plasma NF-L levels (Domingues et al., 2019; Rathbone et al., 2018; Spanier et al., 2019). NF-L is a biomarker of neurodegenerative processes in the CNS and neuronal damage, and its plasma levels are elevated in delirium and neurodegenerative diseases (Ballweg et al., 2021; Casey et al., 2020; Maes et al., 2022b; Yuan and Nixon, 2016).

Moreover, MDD patients have elevated cerebro-spinal fluid (CSF) levels of glial fibrillary acidic protein (GFAP) (Michel et al., 2021), an intermediate filament in the cytoskeleton of astrocytes that is involved in cell communication, cell structure, and astrocyte function (Maes, 2022; Nedergaard et al., 2003; Wang et al., 2017). GFAP expression is increased in response to CNS trauma and reactive astrogliosis, and GFAP may be translocated into the peripheral circulation through BBB disruption (Hol and Pekny, 2015; Yang and Wang, 2015). Therefore, elevated serum GFAP is a biomarker for neurotoxicity, neuronal and glial cell damage (Yang and Wang, 2015).

Enhancement in tau Positron Emission Tomography imaging is detected in people without cognitive abnormalities who are depressed (Babulal et al., 2020). Tau controls and stabilizes microtubule dynamics, axonal transport, and the maintenance of normal synapses (Caamaño-Moreno and Gargini, 2022; Wang et al., 2022) and tau aggregates are connected with neuronal death, Alzheimer’s disease development, and cognitive impairment (Lecordier et al., 2021; Xia et al., 2017). Moreover, P-tau217 is a biomarker for Alzheimer’s disease (Cullen et al., 2021) and may distinguish Alzheimer’s disease from other neurodegenerative disorders (Janelidze et al., 2020; Palmqvist et al., 2020).

In MDD, increasing insulin resistance (IR) is another negative consequence of IRS activation and increased nitro-oxidative toxicity (Morelli et al., 2021). A substantial proportion of depressed people have elevated IR (Hamer et al., 2019), while individuals with type 2 diabetes have approximately double the incidence of MDD (Wang et al., 2019). Many of the neurodegenerative alterations seen in Alzheimer’s disease may be attributable to insulin signaling dysfunctions (De la Monte, 2017). Even in non-diabetic Alzheimer’s patients, IR is related to decreased grey matter volume and cerebral glucose metabolic rate in the hippocampus (Femminella et al., 2021). There is also evidence that dysfunctional calcium signaling may be one of the key processes in neurodegenerative processes and disorders (Bezprozvanny, 2010) and that magnesium deficiency may contribute to neurodegenerative processes (Hlásný, 2020). This is important because the lowered levels of calcium and magnesium in MDD are associated with IRS activation (Al-Dujaili et al., 2019; Al-Hakeim et al., 2022b).

Nevertheless, GFAPR, NF-L, GFAP and P-tau217 were not measured in association with inflammatory and IR markers, calcium, magnesium and the physio-affective phenome of MDD consisting of depressive, anxiety, fatigue and physiosomatic symptoms. Hence, the current study was conducted to examine those proteins in association with CRP, IR, calcium and magnesium in relation to the severity of the physio-affective phenome of MDD.

## Subjects and Methods

### Participants

We recruited 94 MDD patients aged 24-63 years and 47 age-matched healthy subjects as a control group for the current study. Subjects were enrolled between December 2021 and February 2022 at “The Psychiatry Unit,” Al-Hakeem General Hospital, Najaf Governorate, Iraq. The diagnosis was made using criteria from the Diagnostic and Statistical Manual of Mental Disorders, 5th edition (APA, 2013). We excluded patients and controls with a) systemic diseases such as autoimmune disorders, type 1 diabetes mellitus, inflammatory bowel disease, COPD, and chronic kidney disease; b) neurodegenerative or neuroinflammatory disease such as multiple sclerosis, stroke, and Alzheimer’s and Parkinson’s disease; and c) with axis I diagnoses such as psycho-organic disorders, bipolar disorder, generalized anxiety disorders, post-traumatic stress disorder, schizophrenia, OCD, and substance abuse. None of the subjects engaged in regular light sports or heavy exercise. Normal controls were recruited from the same catchment area as staff or their friends and family members, or friends of patients. Controls were excluded for a lifetime history of affective disoders, including MDD, dysthymia, and cyclothymia and a positive family history of depressive disorder. Pregnant and lactating women were excluded from participating in the current study.

Following approval from the ethics committee (IRB) of the College of Science, University of Kufa, Iraq (Document # 5612 / 2021), written informed consent was obtained from all participants in accordance with the guidelines outlined in the current version of the Declaration of Helsinki. The Body mass index (BMI) was assessed on the same day as the clinical interview as body weight in kg/length in m^2^.

### Clinical measurements

The Beck-Depression Inventory (BDI-II) total score (Beck et al., 1996) was used to determine the severity of depression. On the same day, the same senior psychiatrist assessed sociodemographic data, clinical data using a semi-structured interview, and the Fibromyalgia and Chronic Fatigue Syndrome (CFS) Rating (FibroFatigue or FF) scale to measure the severity of CFS-like symptoms (Zachrisson et al., 2002). The severity of anxiety was measured using the Hamilton Anxiety Rating Scale (HAM-A) (Hamilton, 1959). The total sums on these different rating scales were used as overall severity indices of depression, anxiety and physiosomatic symptoms. Moreover, we computed different symptom domains to score key depressive, anxiety and physiosomatic symptoms. Thus, we calculated the “pure BDI” subdomain as the sum of sadness, discouraged about the future, feeling a failure, dissatisfaction, feeling guilty, feeling punished, disappointed in self, critical of self, suicidal ideation, crying, loss of interest, difficulties with decisions, and work inhibition (thus excluding the physio-somatic BDI-II symptoms). The “pure FF” score was computed as the sum of muscular pain, muscle tension, fatigue, autonomous symptoms, gastrointestinal symptoms, headache and a flu-like malaise (thus excluding the affective and cognitive symptoms of the FF scale). Furthermore, we computed “pure HAMA anxiety” and “physiosom HAMA” subscores. The first was computed as sum of sad mood, feelings of guilt, suicidal thoughts, and loss of interest; and the physiosom HAMA as the sum of somatic anxiety, gastrointestinal (GIS) anxiety, genitourinary anxiety, and hypochondriasis. Finally, the “physio-affective phenome” of MDD was computed as the first latent vector extracted from the pure BDI, pure FF, pure HAMA an physiosom HAMA scores. The DSM-5 criteria were used to diagnose tobacco use disorder (TUD). Body mass index (BMI) was computed as weight in kilograms divided by body length in meters squared.

### Assays

In the early morning hours, five milliliter of venous blood was drawn from fasting patients and controls utilizing disposable needles and plastic syringes. The samples were transferred into a clean plain tube. Blood was left at room temperature for 15 min for clotting, then centrifuged 3000 rpm for 10 min, and then serum was separated and transported into Eppendorf tubes to be stored at −80 °C until thawed for assay. The C-Reactive Protein (CRP) latex slide test (Spinreact^®^, Barcelona, Spain) was used for CRP assays in human serum. Commercial ELISA sandwich kits provided by Sunlong Biotech Co., Ltd (Zhejiang, China), and Melsin Medical Co. (Jilin, China) were used to measure GFAP, NF-L, P-tau217, and PDGFRβ. Commercial ELISA sandwich kits were used to measure serum insulin (DRG^®^ International Inc., USA). Calcium, magnesium and glucose levels in serum were measured spectrophotometrically using a ready to use kit supplied by Spinreact^®^ (Barcelona, Spain). We followed all the procedures exactly according to the manufacturer’s instructions. The insulin sensitivity percentage (HOMA%S), percentage of beta-cell function (HOMA%B) and insulin resistance value (HOMA2IR) were measured from fasting insulin and serum glucose by HOMA2 Calculator downloaded from (https://wwwdtuoxacuk/homacalculator/). The sensitivities of the GFAP, NF-L, P-tau217 and PDGFRβ assays was 0.1 pg/ml and the intra-assay coefficient of variation (CV) for those assays was <10.0% (precision within-assay). The sensitivity of the insulin assay was 1.76 µIU/ml (12.22pM). and the CV% was 2.6%.

### Statistical analysis

Analysis of variance (ANOVA) was used to check if there were any differences in scale variables between groups, and the analysis of contingency tables (χ^2^-test) was employed to assess the associations between nominal variables. We used correlation matrices based on Pearson’s product-moment or Spearman’s rank-order correlation coefficients to assess the relationships between biomarkers and clinical scores. While adjusting for possible intervening variables such as sex, age, TUD, BMI, and education, multivariate general linear model (GLM) analysis was used to check the associations between biomarkers and diagnostic groups. To determine the associations between diagnostic classes and biomarkers, between-subject effects were tested, and effect sizes were estimated using partial eta-squared values. We calculated GLM-generated estimated marginal mean (SE) values and performed protected pairwise comparisons between treatment means. Using clinical and biomarker levels as explanatory variables in an automatic stepwise method with a p-to-enter of 0.05 and p-to-remove of 0.06, multiple regression analysis was used to delineate the significant biomarkers predicting the physio-somatic scores. Using VIF and tolerance values, all results were checked for multicollinearity. Statistical tests were two-tailed, and statistical significance was determined using a p-value of 0.05. IBM SPSS windows version 25, 2017 was used for all statistical analyses.

SmartPLS analysis (Ringle et al., 2015) was used to investigate the causal links between CRP, calcium, the neuroaxis, PDGFRβ and HOMA2-IR (and socio-demographic data) and the physio-affective phenome of MDD. The latter variable was a latent vector derived from pure BDI, FF and HAMA and the physiosom HAMA scores. The neuroaxis was entered as a latant vector extracted from GFAP, NF-L, P-Tau217 and their composite, whilst all other input variables were entered as single indicators. Only when the outer and inner models fulfilled the following quality criteria was a complete PLS analysis performed: 1) the model fit is <0.08 in terms of standardized root squared residual (SRMR) values; 2) the output latent vector has strong construct and convergence validity, as shown by rho A >0.8, Cronbach’s alpha >0.7, composite reliability >0.7, and average variance extracted (AVE) greater than 0.5, 3) all loadings on the extracted latent vector are more than 0.6 at p<0.001, 4) Blindfolding shows that the construct’s cross-validated redundancy is sufficient; 5) Confirmatory Tetrad Analysis (CTA) shows that the latent vector extracted from the rating scale scores is not mis-specified as a reflective model; 6) PLS Predict shows that the model’s prediction performance is satisfactory. If all of the model quality data meets the predefined requirements, we perform a complete PLS-SEM path analysis with 5,000 bootstrap samples, calculating the path coefficients (with p-values), as well as specific and total indirect (mediated) effects and total effects.

To interpret the results of the study, we performed annotation and enrichment analysis using the differentially expressed proteins (DEPs) identified in this study (gene symbols, namely NEFL (neurofilament light polypeptide), GFAP, MAPT (mictotubule-associated protein tau) and PDGFRB (platelet-derived growth factor receptor beta) as seed genes. The latter were enlarged with known protein interactions from GOnet (https://tools.dice-database.org/Gonet/) and Enrichr, a gene list enrichment analysis tool (https://maayanlab.cloud/Enrichr/), and their pathway and function enrichment scores were calculated. We searched the Enrichr network for GO biological processes (assemblies of molecular activities in pathways) and GO cellular components (location of proteins) (www.geneontology.org) and visualized the results as Appyter bar graphs. Furthermore, GOnet analysis was used to create interactive graphs that included the four selected DEPs and GO terms. We utilize p-values that have been adjusted for the false discovery rate (FDR).

## Results

### 1. Socio-demographic data

**Table 1** shows the socio-demographic and clinical data in healthy controls and MDD patients divided into two groups, namely those with severe depression (defined as a total BDI score ≥ 34) and those with moderate MDD (BDI < 34). There were no significant differences in age, BMI, marital status ratio, TUD, sex ratio, and rural/urban ratio among the three study groups. The severe-MDD group showed a trend towards lower education as compared with the control group. There is a significant decrease in the employment ratio in patient groups compared with the control group, with the lowest ratio in the severe MDD. All rating scale scores and their subdomains were significantly different between the three groups and increased from controls to moderate MDD to severe MDD, although the pure HAMA score was significantly higher in patients than controls.

**Table 1.**
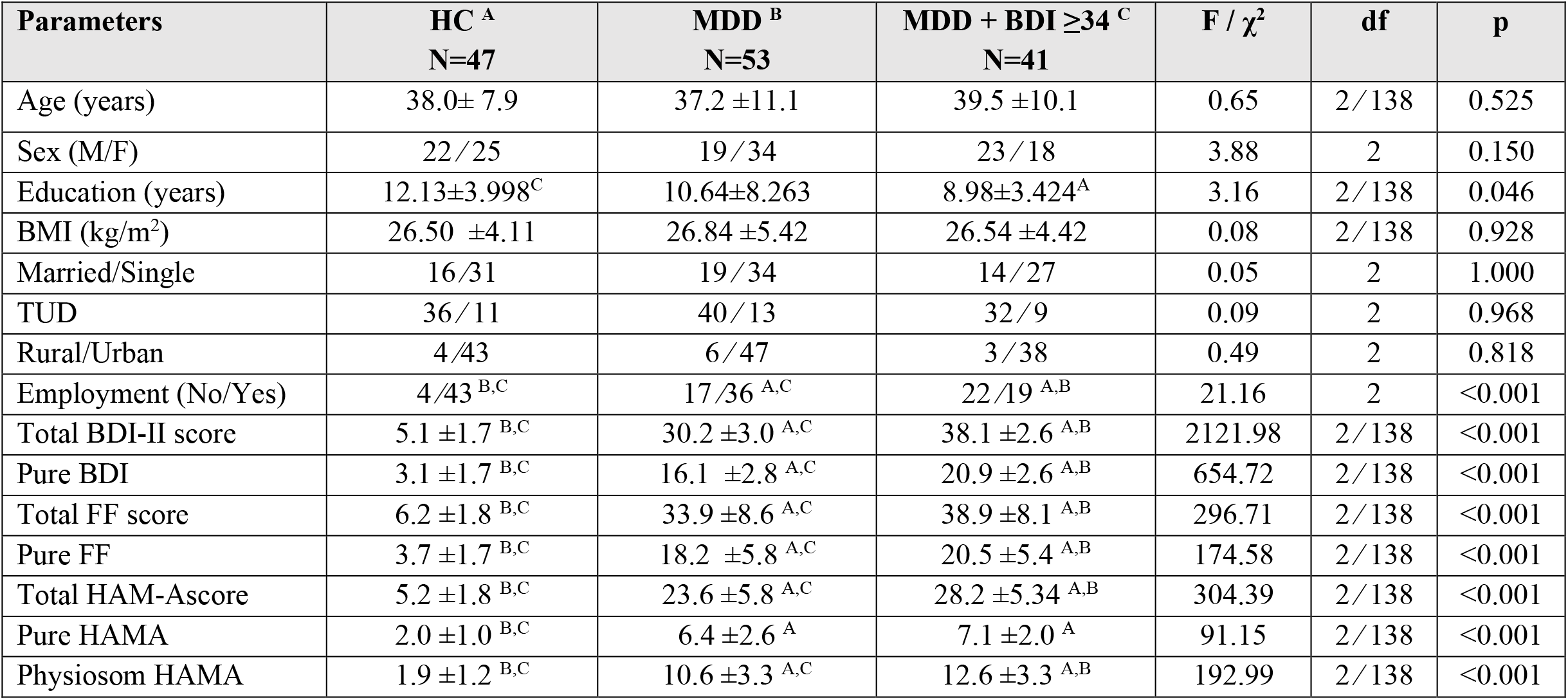
Demographic and clinical data of the participants in this study, namely healthy controls (HC) and major depressed patients (MDD) divided into two groups based on the Beck-Depression Inventory (BDI) scores ≥ or < than 34. Results are shown as mean ±standard deviation (SD) or as ratios. F: results of analyses of variance, χ^2^ : results of contingency analysis; ^A, B, C^: Pairwise comparison among group means. TUD: Tobacco use disorder, BMI: body mass index, HAM-A: Hamilton Anxiety Rating Scale score, BDI-II: Beck Depression Inventory, Pure BDI: key depressive symptoms of the BDI, Pure FF: key physiosomatic symptoms of the Fibromyalgia and Chronic Fatigue Syndrome Rating (FF) scale, Pure HAMA: key anxiety symptoms of the Hamilton Anxiety Rating Scale (HAMA) score, Physiosom HAMA: key phyio-somatic symptoms of the HAMA score.

### 2. Biomarkers in Moderate-MDD and Severe-MDD

**Table 2** shows the results of multivariate GLM analysis which examines the association between eleven biomarkers (GFAP, NF-L, P-tau217, CRP, PDGFRβ, magnesium, calcium, HOMA2 %S, HOMA2 IR, FBG, and insulin) and diagnosis (controls versus moderate MDD versus severe MDD) while adjusting for age, sex, BMI, and TUD. We found that there was a significant association between diagnosis and the levels of the variables, with an effect size of 0.366, while there were no significant effects of the covariates. Tests for between-subject effect analyses showed that there were significant associations between diagnosis and (in descending order of importance) NF-L (effect size=partial η^2^ = 0.406), GFAP (0.246), calcium (0.214), CRP (0.171), P-tau217 (0.143), PDGFRβ (0.139), HOMA2 %S (0.134), insulin (0.114), and HOMA2-IR (partial η^2^=0.111).

**Table 2:** Results of multivariate GLM analysis with all biomarkers as dependent variables and covariates including BDI subgroups as explanantrory variables. CRP: C-reactive protein, HOMA2IR: homeostasis model assessment of insulin resistance, HOMA2%S: homeostasis model assessment of insulin sensitivity percentage, PDGFRβ: Platelet-derived growth factor-receptor beta, NF-L neurofilament light chain, PTau217: phosphorylated tau-217 protein, and GFAP: Glial fibrillary acidic protein.

| Tests | Dependent variables | Explanatory variables | F | df | p | Partial $\eta^2$ |
| --- | --- | --- | --- | --- | --- | --- |
| **Multivariate** | All biomarkers | HC_MDD groups | 6.57 | 22/250 | <0.001 | 0.366 |
|  |  | Age | 0.97 | 11/124 | 0.480 | 0.079 |
|  |  | BMI | 1.28 | 11/124 | 0.246 | 0.102 |
|  |  | TUD | 0.69 | 11/124 | 0.746 | 0.058 |
|  |  | Sex | 0.99 | 11/124 | 0.461 | 0.081 |
| **Between-subject effects** | GFAP | HC_MDD groups | 21.86 | 2/134 | <0.001 | 0.246 |
|  | NF-L | HC_MDD groups | 45.70 | 2/134 | <0.001 | 0.406 |
|  | P-tau217 | HC_MDD groups | 11.14 | 2/134 | <0.001 | 0.143 |
|  | CRP | HC_MDD groups | 13.80 | 2/134 | <0.001 | 0.171 |
| | PDGFRβ | HC_MDD groups | 10.78 | 2/134 | <0.001 | 0.139 |
|  | Magnesium | HC_MDD groups | 0.33 | 2/134 | 0.723 | 0.005 |
|  | Calcium | HC_MDD groups | 18.24 | 2/134 | <0.001 | 0.214 |
|  | HOMA2 %S | HC_MDD groups | 10.41 | 2/134 | <0.001 | 0.134 |
|  | HOMA2-IR | HC_MDD groups | 8.39 | 2/134 | <0.001 | 0.111 |
|  | Fasting Blood Glucose | HC_MDD groups | 2.08 | 2/134 | 0.129 | 0.030 |
|  | Insulin | HC_MDD groups | 8.64 | 2/134 | <0.001 | 0.114 |

CRP: C-reactive protein, HOMA2IR: homeostasis model assessment of insulin resistance, HOMA2%S: homeostasis model assessment of insulin sensitivity percentage, PDGFRβ : Platelet-derived growth factor-receptor beta, NF-L neurofilament light chain, PTau217: phosphorylated tau-217 protein, and GFAP: Glial fibrillary acidic protein.

**Table 3** shows the model-generated estimated mean (± SE) biomarker values in the three study groups. P-tau217, PDGFRβ, HOMA2IR, and insulin were significantly increased in MDD patients in comparison with controls. GFAP and NF-L showed significant differences between the three study groups and increased from controls to moderate MDD to severe MDD. HOMA2S% and calcium were significantly decreased in both patient groups compared with controls. FBG was significantly higher in moderate MDD compared with the control group. CRP was significantly increased in the severe MDD group compared with the moderate MDD and control group.

**Table 3.** Model-generated estimated marginal mean values of the measured biomarkers in healthy controls (HC) and major depressed patients (MDD) divided into two groups based on the Beck-Depression Inventory (BDI) scores ^A, B, C^: Pairwise comparison among group mean values; CRP: C-reactive protein, FBG: fasting blood glucose, HOMA2-IR: homeostasis model assessment of insulin resistance, HOMA2%S: homeostasis model assessment of insulin sensitivity percentage, PDGFRbeta: Platelet-derived growth factor receptor beta, NF-L neurofilament light chain, P-tau217: phosphorylated tau-217 protein, and GFAP: Glial fibrillary acidic protein.

| Parameter | HC <sup>A</sup><br>N=47 | MDD BDI < 34 <sup>B</sup><br>N=53 | MDD BDI ≥ 34 <sup>C</sup><br>N=41 |
| --- | --- | --- | --- |
| GFAP (pg/mL) | 116.7 ±11.2 <sup>B,C</sup> | 169.9 ±10.6 <sup>A,C</sup> | 225.4 ±12.1 <sup>A,B</sup> |
| NF-L (pg/mL) | 11.1 ±1.19 <sup>B,C</sup> | 23.0 ±1.13 <sup>A,C</sup> | 26.72 ±1.29 <sup>A,B</sup> |
| P-tau 217 (pg/mL) | 1.12 ±0.13 <sup>B,C</sup> | 1.66 ±0.12 <sup>A</sup> | 1.99 ±0.14 <sup>A</sup> |
| CRP (mg/L) | 2.74 ±0.86 <sup>C</sup> | 4.94 ±0.82 <sup>C</sup> | 9.35 ±0.94 <sup>A,B</sup> |
| PDGFRβ (ng/mL) | 4.03 ±0.29 <sup>B,C</sup> | 5.74 ±0.28 <sup>A</sup> | 5.63 ±0.32 <sup>A</sup> |
| Magnesium (mM) | 1.03 ±0.03) | 1.01 ±0.03 | 1.01 ±0.03 |
| Calcium (mM) | 2.39 ±0.02 <sup>B,C</sup> | 2.25 ±0.02 <sup>A</sup> | 2.22 ±0.02 <sup>A</sup> |
| HOMA2 S% | 91.73 ±2.93 <sup>B,C</sup> | 77.41 ±2.79 <sup>A</sup> | 73.46 ±3.18 <sup>A</sup> |
| HOMA2-IR | 1.15 ±0.057 <sup>B,C</sup> | 1.41 ±0.05 <sup>A</sup> | 1.45 ±0.06 <sup>A</sup> |
| FBG (mM) | 5.27 ±0.11 <sup>B</sup> | 5.58 ±0.11 <sup>A</sup> | 5.49 ±0.12 |
| Insulin (pM) | 60.6 ±2.8 <sup>B,C</sup> | 73.82 ±2.70 <sup>A</sup> | 76.27 ±3.07 <sup>A</sup> |

### 3. Multiple regression analysis with psychiatric rating scale scores as dependent variables

**Table 4** shows the results of different stepwise multiple regression analyses with the psychiatric rating scale scores reflecting depression, anxiety and physiosomatic symptoms as dependent variables, while allowing for the effects of age, sex, and education. Regression #1 shows that 62.9 % of the variance in the Pure BDI score was explained by the regression on NF-L, GFAP, P-tau217, and PDGFRβ, and inversly with calcium and HOMA2%S. **Figure 1** shows the partial regression plot of the pure BDI score on NF-L after considering the effects of CRP, HOMA2 %S and calcium. Regression #2 shows that 52.7% of the variance in the Pure FF score can be explained by the regression on NF-L, P-tau217, PDGFRβ, and GFAP (positively) and calcium and HOMA2%S (inversely). Regression #3 shows that 47.5% of the variance in the Pure HAMA score was explained by NF-L, HOMA2 IR, PDGFRβ, and P-tau217.

**Figure 1.**
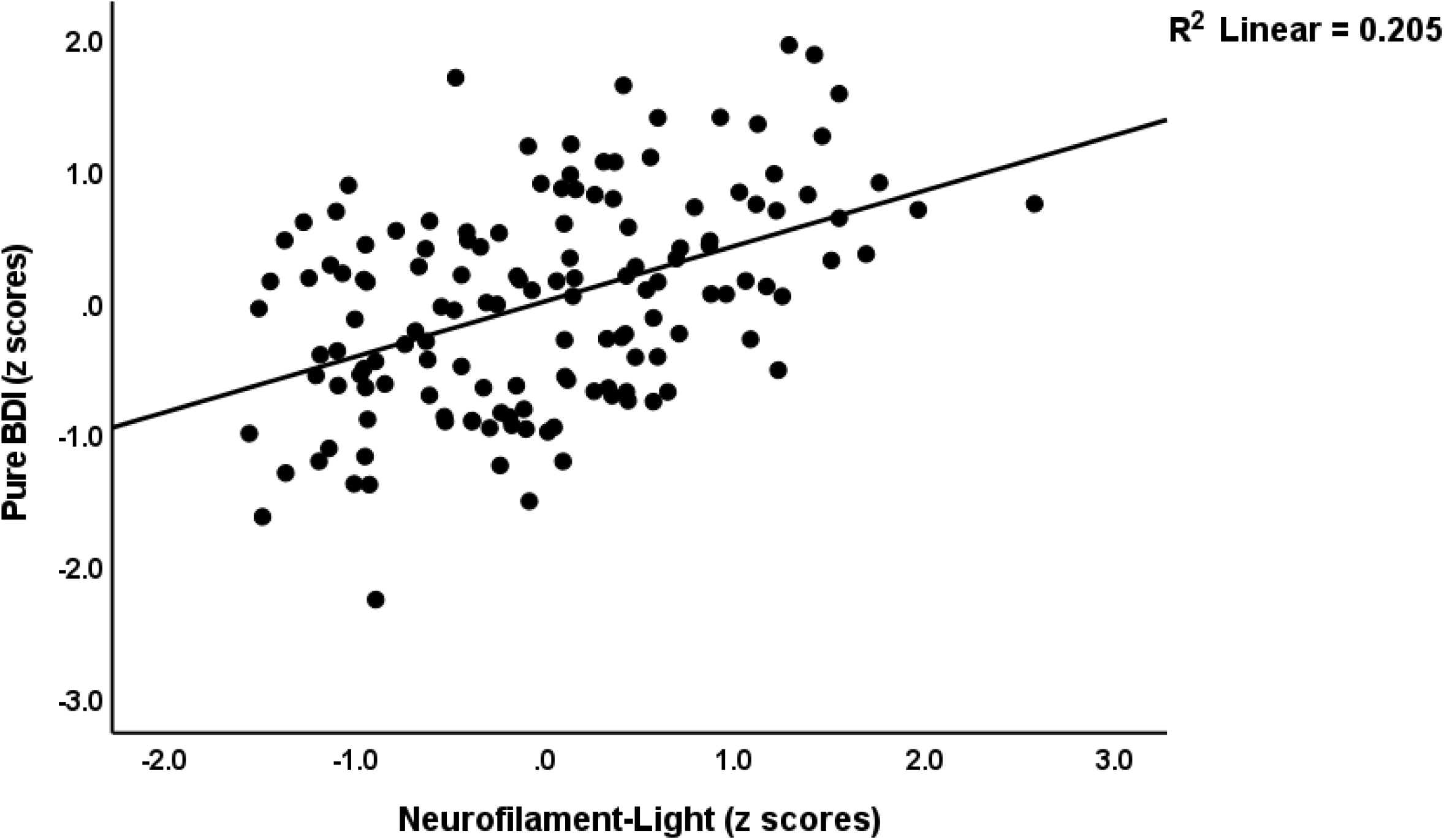
Partial regression of the pure depressive symptoms of the Beck Depression Inventory on serum neurofilament light concentrations

**Table 4.** Results of multiple regression analyses with physio-affective symptom domain scores reflecting depression, anxiety and physio-somatic symptoms as dependent variables. Physio-affective phenome: first factor extracted from Pure BDI, Pure FF, Pure HAMA and Physiosom HAMA, CRP: C-reactive protein, FBG: fasting blood glucose, HOMA2-IR: homeostasis model assessment of insulin resistance, HOMA2 %S: homeostasis model assessment of insulin sensitivity percentage, PDGFRbeta: Platelet-derived growth factor-beta, NF-L neurofilament light chain, P-tau217: phosphorylated tau-217 protein, GFAP: Glial fibrillary acidic protein; neuroaxis index: first principal componenet extracted from GFAP, GFAP and NF-L HAM-A: Hamilton Anxiety Rating Scale score, BDI-II: Beck Depression Inventory, Pure BDI: key depressive symptoms of the BDI, Pure FF: key physiosomatic symptoms of the Fibromyalgia and Chronic Fatigue Syndrome Rating (FF) scale, Pure HAMA: key anxiety symptoms of the Hamilton Anxiety Rating Scale (HAMA) score, Physiosom HAMA: key phyio-somatic symptoms of the HAMA score.

| Regression | Explanatory variables | $\beta$ | t | p | F<sub>model</sub> | df | p | R<sup>2</sup> |
| --- | --- | --- | --- | --- | --- | --- | --- | --- |
| #1. Pure BDI | Model |  |  |  | 37.51 | 6/133 | <0.001 | 0.629 |
|  | NF-L | 0.365 | 5.97 | <0.001 |  |  |  |  |
|  | GFAP | 0.260 | 4.58 | <0.001 |  |  |  |  |
|  | P-tau217 | 0.223 | 3.85 | <0.001 |  |  |  |  |
|  | Calcium | -0.199 | -3.32 | 0.001 |  |  |  |  |
| | PDGFRβ | 0.151 | 2.74 | 0.007 | | | | |
|  | HOMA2 %S | -0.133 | -2.26 | 0.025 |  |  |  |  |
| #2. Pure FF | Model |  |  |  | 24.88 | 6/134 | <0.001 | 0.527 |
|  | NF-L | 0.298 | 4.35 | <0.001 |  |  |  |  |
|  | HOMA2 %S | -0.255 | -3.89 | <0.001 |  |  |  |  |
|  | P-tau217 | 0.205 | 3.13 | 0.002 |  |  |  |  |
| | PDGFRβ | 0.177 | 2.87 | 0.005 | | | | |
|  | Calcium | -0.162 | -2.41 | 0.018 |  |  |  |  |
|  | GFAP | 0.136 | 2.13 | 0.035 |  |  |  |  |
| #3. Pure HAMA | Model |  |  |  | 25.88 | 4/135 | <0.001 | 0.475 |
|  | NF-L | 0.337 | 4.90 | <0.001 |  |  |  |  |
|  | HOMA2-IR | 0.250 | 3.59 | <0.001 |  |  |  |  |
| | PDGFRβ | 0.254 | 3.80 | <0.001 | | | | |
|  | P-tau217 | 0.198 | 2.91 | 0.004 |  |  |  |  |
| #4. Physiosom HAMA | Model |  |  |  | 23.17 | 6/134 | <0.001 | 0.509 |
|  | NF-L | 0.320 | 4.59 | <0.001 |  |  |  |  |
|  | P-tau217 | 0.212 | 3.19 | 0.002 |  |  |  |  |
|  | Calcium | -0.158 | -2.30 | 0.023 |  |  |  |  |
|  | GFAP | 0.181 | 2.79 | 0.006 |  |  |  |  |
| | PDGFRβ | 0.168 | 2.66 | 0.009 | | | | |
|  | HOMA2 %S | -0.176 | -2.63 | 0.010 |  |  |  |  |
| #5. Physio-affective phenome score | Model |  |  |  | 33.80 | 6/133 | <0.001 | 0.604 |
|  | NF-L | 0.343 | 5.45 | <0.001 |  |  |  |  |
|  | P-tau217 | 0.223 | 3.72 | <0.001 |  |  |  |  |
|  | Calcium | -0.167 | -2.70 | 0.008 |  |  |  |  |
| | PDGFRβ | 0.188 | 3.30 | 0.001 | | | | |
|  | HOMA2%S | -0.210 | -3.46 | 0.001 |  |  |  |  |
|  | GFAP | 0.186 | 3.17 | 0.002 |  |  |  |  |
| #6. Physio-affective<br>phenome score | **Model** |  |  |  | 19.47 | 4/136 | <0.001 | 0.587 |
|  | Neuroaxis index | 0.490 | 8.21 | <0.001 |  |  |  |  |
|  | Calcium | -0.217 | -3.73 | <0.001 |  |  |  |  |
|  | HOMA2-IR | 0.230 | 3.90 | <0.001 |  |  |  |  |
| | PDGFRβ | 0.196 | 3.43 | <0.001 | | | | |

Regression #4 shows that 50.9% of the variance in the physiosom HAMA score can be explained by the regression on NF-L, P-tau217, GFAP, and PDGFRβ (all positively), and calcium and HOMA2%S (all inversely). In regression #5, 60.4% of the variance in the physio-affective phenome (computed as the first factor extracted from pure BDI, pure FF, pure HAMA and physiosom_HAMA) can be explained by the regression on NF-L, P-tau217, PDGFRβ, and GFAP (all positively), and HOMA2 %S and calcium (both inversely associated). We have computed a neuroaxis index by extracting the first principal component from GFAP, P-tau217 and NF-L. This first principal component explained 52.24% of the variance and all three variables loaded highly (>0.611) (PDGFRβ did not load highly enough to be included in this component). Regression #6 shows that 58.7% of the variance in the physio-affective phenome was explained by the neuroaxis index, calcium, HOMA2-IR and PDGFRβ. **Figure 2** shows the partial regression plot of the physio-affective phenome on the neuroaxis index after controlling for calcium, HOMA2-IR and PDGFRβ.

**Figure 2.**
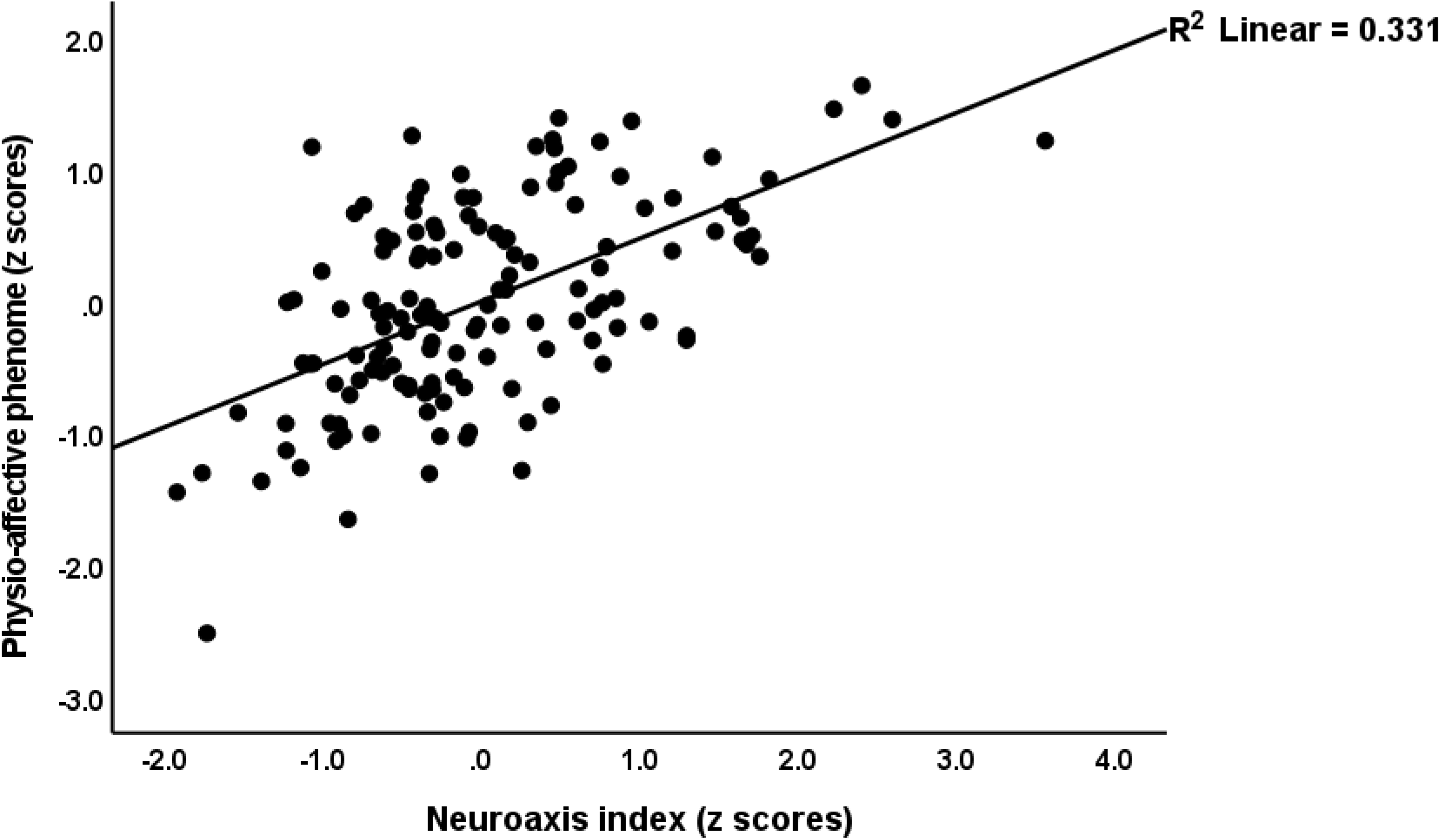
Partial regression of the physio-affective phenome score (which combines fatigue, physio-somatic and pure depressive and anxiety scores) on the neuroaxis composite score

### 4. Multiple regression analysis with biomarkers as dependent variables

**Table 5** shows the results of multiple regression analyses with the neuroaxis biomarkers as dependent variables. Regression #1 shows that 27.7% of the variance in the neuroaxis index was explained by the regression of CRP and IR (both positively correlated). **Figure 3** shows the partial regression of the neuroaxis index on CRP. Regression #2 shows that a considerable part of the variance in NF-L (26.1%) was explained by the regression on calcium (negatively), CRP (positively) and being female. We found that CRP explained 10.3% of the variance in the GFAP levels (see Regression #3). Regression #4 shows that 17.5% of the variance in P-Tau217 was explained by the combined effects of CRP and HOMA2-IR. Regression #5 shows that 39.9% of the variance in PDGFRβ was explained by calcium (negatively associated).

**Figure 3.**
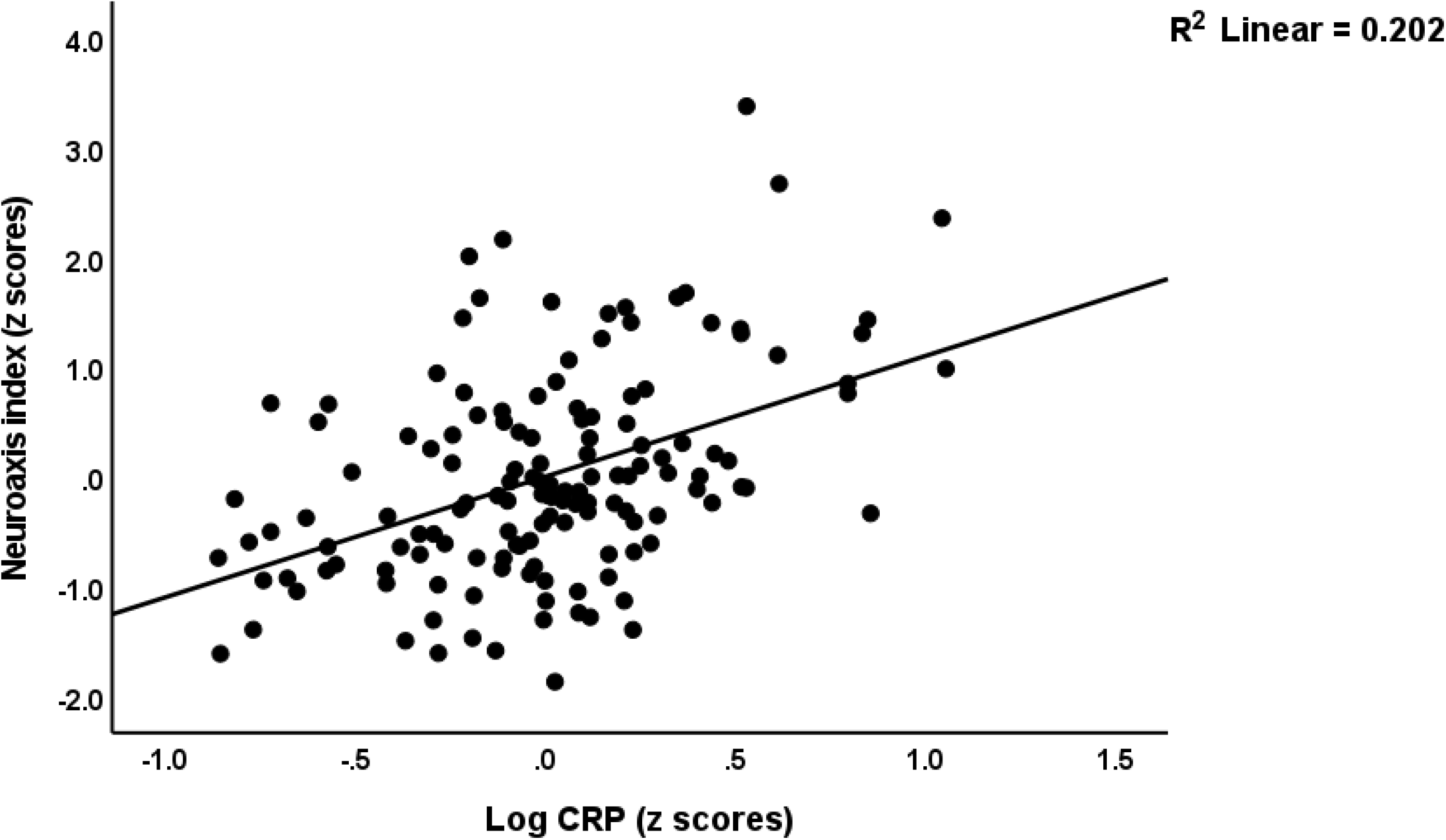
Partial regression of the neuroaxis index score on serum C-reactive protein (in logarithmic and z transformation)

**Table 5.** Results of multiple regression analysis with biomarkers as dependent variables. Neuroaxis index: first principal component extracted from NF-L (neurofilament light chain), P-tau217: (phosphorylated tau-217 protein),and GFAP (glial fibrillary acidic protein); PDGFRbeta: Platelet-derived growth factor-beta CRP: C-reactive protein, FBG: fasting plasma glucose, HOMA2-IR: homeostasis model assessment of insulin resistance, HOMA2 %S: homeostasis model assessment of insulin sensitivity percentage,

| Regression | Explanatory variables | $\beta$ | t | p | F<sub>model</sub> | df | p | R<sup>2</sup> |
| --- | --- | --- | --- | --- | --- | --- | --- | --- |
| #1. Neuroaxis index | **Model** |  |  |  | 26.41 | 2/138 | <0.001 | 0.277 |
|  | CRP | 0.435 | 5.91 | <0.001 |  |  |  |  |
|  | HOMA2-IR | 0.226 | 3.07 | 0.003 |  |  |  |  |
| #2. NF-L | **Model** |  |  |  | 16.15 | 3/137 | <0.001 | 0.261 |
|  | Calcium | -0.353 | -4.69 | <0.001 |  |  |  |  |
|  | CRP | 0.286 | 3.77 | <0.001 |  |  |  |  |
|  | Male Sex | -0.153 | -2.07 | 0.040 |  |  |  |  |
| #3. GFAP | **Model** |  |  |  | 15.94 | 3/139 | <0.001 | 0.103 |
|  | CRP | 0.321 | 3.99 | <0.001 |  |  |  |  |
| #4. P-tau217 | **Model** |  |  |  | 14.59 | 2/138 | <0.001 | 0.175 |
|  | CRP | 0.305 | 3.90 | <0.001 |  |  |  |  |
|  | HOMA2 IR | 0.245 | 3.14 | 0.002 |  |  |  |  |
| #5. PDGFRβ | **Model** | | | | 5.21 | 1/139 | 0.024 | 0.036 |
|  | Calcium | -0.190 | -2.28 | 0.024 |  |  |  |  |

#### Results of PLS-SEM path analysis

**Figure 4** shows the results of a complete PLS analysis performed using 5,000 bootstrap samples. The model quality was more than adequate, with an SRMR of 0.041. We observed appropriate construct reliability validity values for the physio-affective phenome with AVE = 0.843; rho A = 0.942; composite reliability = 0.956; and Cronbach alpha = 0.938, while all loadings of the 4 indicators of the physio-affective phenome were greater than 0.87. Moreover, the construct reliability and convergenec of the neuroaxis latent vector were also adequate with AVE = 0.607; rho A = 0.817; composite reliability = 0.857; and Cronbach alpha = 0.770, while all loadings of the indicators were greater than 0.67. According to CTA, both latent vectors were not mis-specified as reflective models and blindfolding revealed an acceptable construct cross-validated redundancy of 0.504 for the physio-affective phenome and 0.160 for the neuroaxis. The construct indicators had positive Q2 predict values, suggesting that the prediction error was less than the naivest benchmark, according to PLSPredict. We found that 61.1% of the variance in the physio-affective phenome is explained by the regression on the neuroaxis index, HOMA2-IR, calcium and PDGFRβ. In addition, 28.9% of the variance in the neuroaxis was predicted by CRP and HOMA2-IR. There were significant specific indiect effects of CRP mediated by the neuroaxis (t=5.37, p<0.001) and calcium, which were partially mediated by HOMA2-IR (t-2.42, p=0.008) and the path from HOMA2-IR to the neuroaxis (t=2.30, p=0.011) on the physio-affective phenome of MDD.

**Figure 4.**
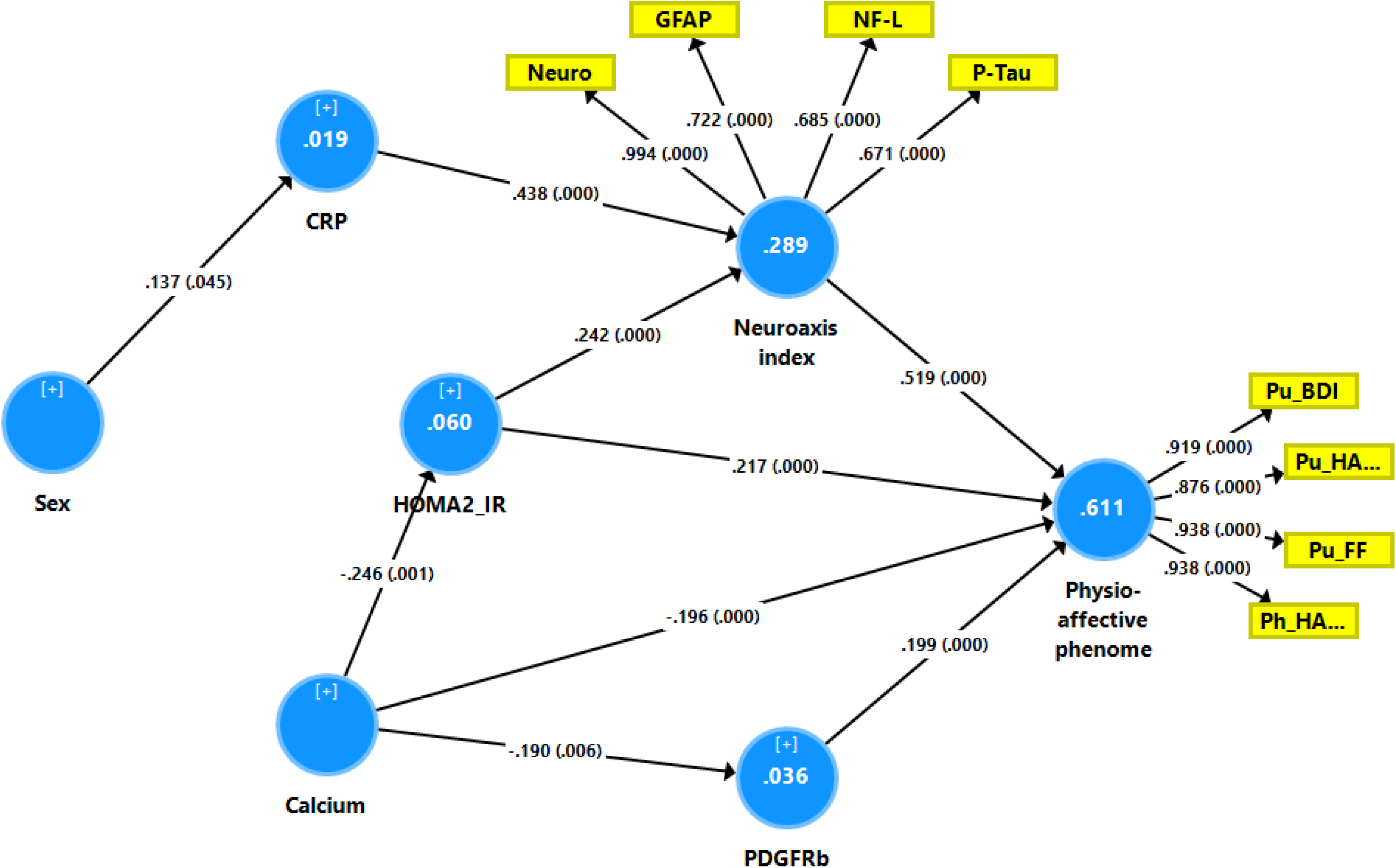
Results of partial least squares (PLS) analysis with the physio-affective phenome as final outcome variable and the neuroaxis index score, the HOMA2-IR index, calcium, platelet-derived growth factor receptor (PDGFR)β, C-reactive protein and sex as explanatory indicators. The physio-affective phenome is entered as a latent vector extracted from 4 symptomatic subdomains, namely pure depression symptoms of the Beck Depression Inventory-II (Pu_BDI), pure anxiety symptoms of the Hamilton Anxiety rating Scale (Pu_HA..), physio-somatic symptoms of the Hamilton Anxiety rating Scale (Ph_HA…), and the pure physio-somatic symptoms of the Fibro-Fatigue Scale (Pu_FF). The neuroaxis index was entered as a latent vector extracted from the serum levels of glial fibrillary acidic protein (GFAP), neurofilament light (NF-L) and phosphorylated tau protein 217 (P-tau217). Non-significant paths and indicators are omitted from the model. The model shows the explained variances (figures in the blue circles), path coefficients (with p values) and the loadings (with p values) of the outer model.

#### Results of annotation and enrichment analysis

**Figure 5** shows the top 10 GO cellular components enriched in the DEP network indicating that glial cell (descendent GO:0097449 or astrocyte projection) and neuron projection, axons, and the skeleton are the most important cellular componenets. **Figure 6** displays the top 10 GO biological processes that were significantly over-represented in the enlarged network, namely axonal transport of mitochondrion, intermediate filament organization, axonal transport, and also regulation of oxygen species metabolic process. **Figure 7** shows the GOnet analysis with the GO terms and the 4 DEPS as seed genes. The GO terms comprised the same functions as those described above, namely cytoskeleton organization and supramolecular fiber organization (p adjusted <1.46E-0.2, comprising the 4 DEPS) and all other terms displayed in the figure (all FDR p adjusted < 1.46 E-2, comprising combination of two DEPS i.e., GFAP and NEFL or MAPT and NEFL).

**Figure 5.**
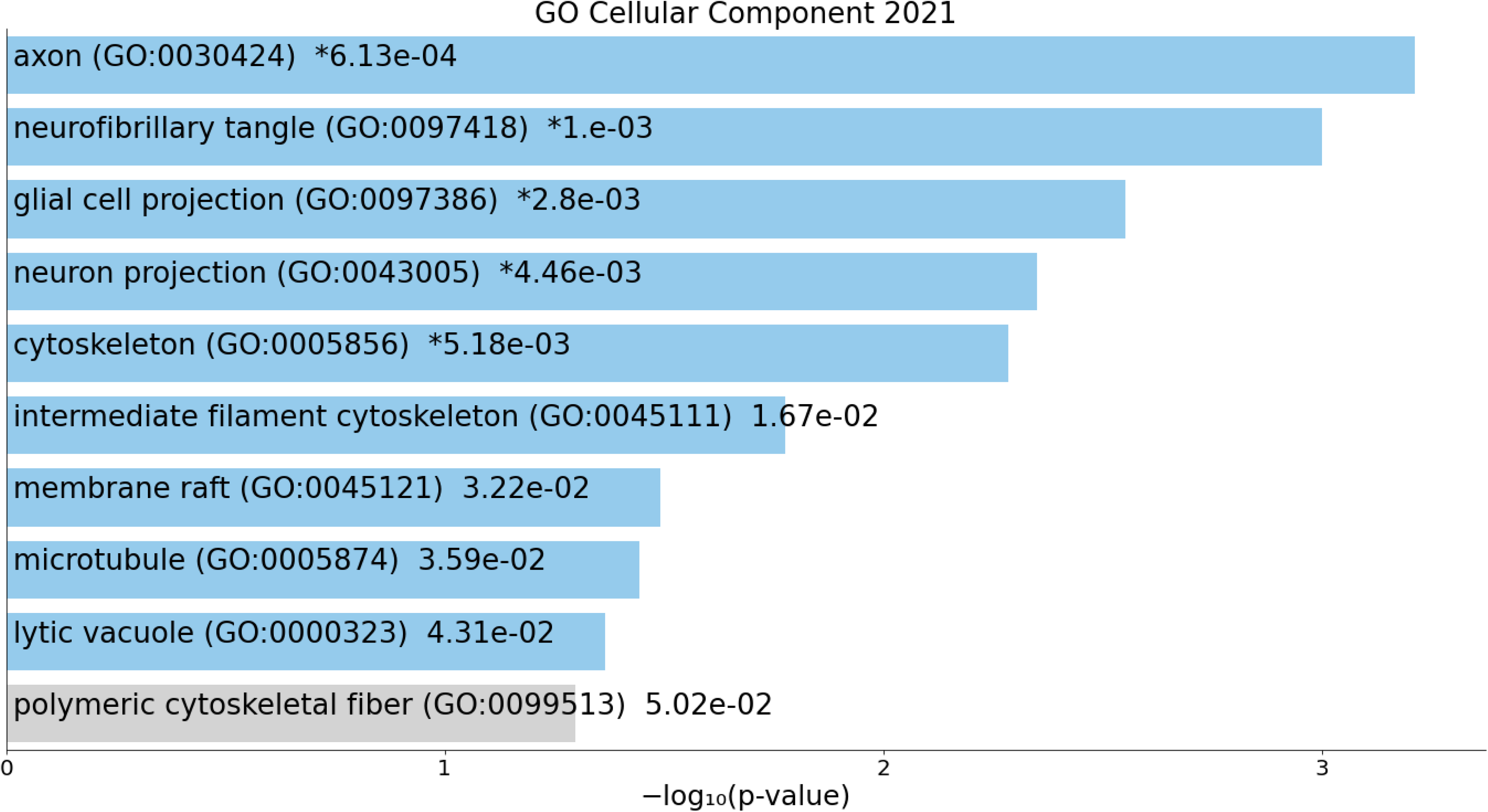
Heatmap of the top 10 enriched GO Cellular Component (2021) terms showing the cell structures that are overexpressed in the enlarged Enrichr (Enrichr (maayanlab.cloud), as accessed 1 July 2022) network constructed using glial fibrillary acidic protein, neurofilament light, and phosphorylated tau protein 217 and platelet-derived growth factor receptor β as seed genes. “Colored bars correspond to terms with significant p-values (<0.05); and the * indicate that the term has a significant adjusted p-value (<0.05)”. The bargraph was made employing Appyters, Appyter (maayanlab.cloud) as accessed 1 July 2022.

**Figure 6.**
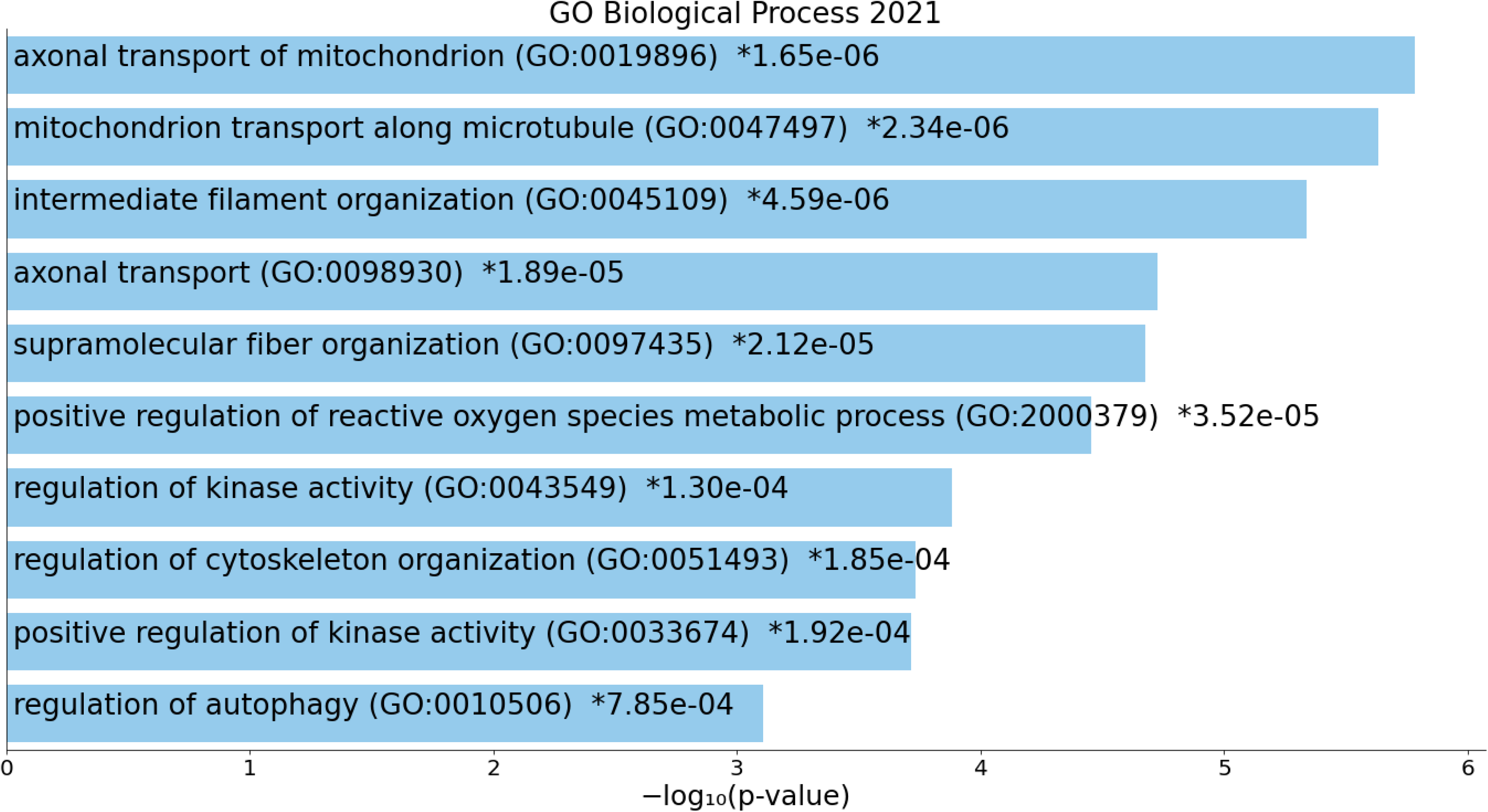
Heatmap of the top 10 enriched GO Biological Process (2021) terms showing the functions that are overexpressed in the enlarged Enrichr (Enrichr (maayanlab.cloud), as accessed 1 July 2022) network constructed using glial fibrillary acidic protein, neurofilament light, phosphorylated tau protein 217 and platelet-derived growth factor receptor β as seed genes. “Colored bars correspond to terms with significant p-values (<0.05); and the * indicate that the term has a significant adjusted p-value (<0.05)”. The bargraph was made employing Appyters, Appyter (maayanlab.cloud) as accessed 1 July 2022.

**Figure 7.**
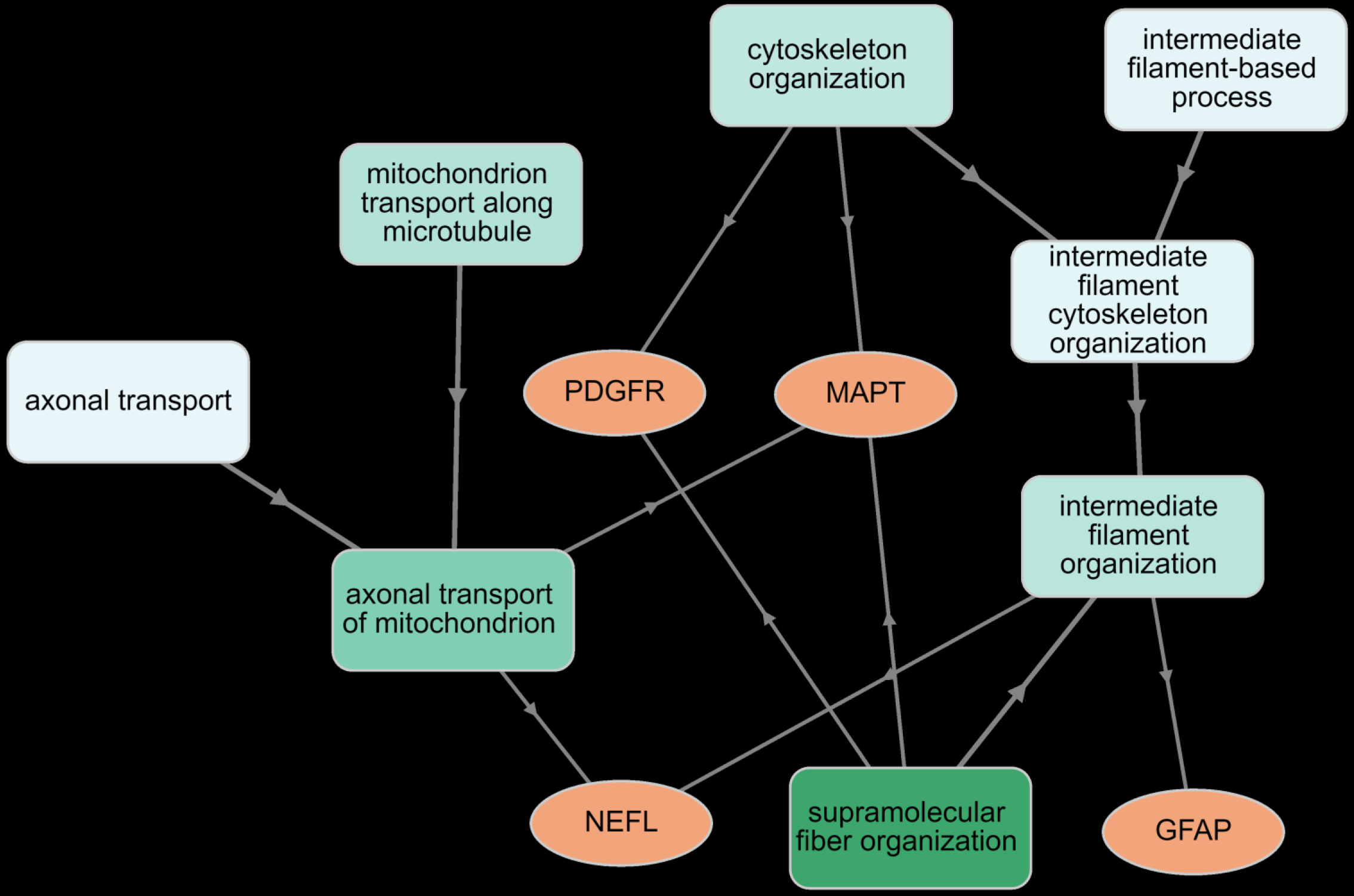
Results of GOnet annotation visualization (Euler layout) showing the hierarchical structure of glial fibrillary acidic protein (GFAP), neurofilament light (NEFL), phosphorylated tau protein 217 (MAPT) and platelet-derived growth factor receptor β (PDGFRB) as seed genes and the corresponding GO terms. This figure shows the seed protein nodes and the top 8 GO terms. The differentially expressed genes are connected with the most specific GO annotations and additionally can be connected to less specific GO functions. All GO terms are significant after p correction for false discovery rate (all < 1.2 E-2).

## Discussion

### The astrocyte and neuron injury biomarkers are increased in MDD

The first major finding of the present study is that serum levels of PDGFR, NF-L, GFAP, and P-tau-217 are higher in MDD patients than in controls. These results expand those of prior papers demonstrating that serum levels of NF-L are higher in MDD and treatment-resistant depression (Chen et al., 2022; Domingues et al., 2019; Jakobsson et al., 2014; Rathbone et al., 2018; Spanier et al., 2019). Other research indicates that the mean CSF and serum concentration of NF-L is higher in individuals with bipolar disorder and other mental illnesses compared to healthy controls (Isgren et al., 2017; Spanier et al., 2019; Tauil et al., 2021). People with MDD have lower cognitive function when their serum NF-L levels are higher (Bavato et al., 2021) and people with post-stroke depression also have higher serum NF-L levels (Zhao et al., 2020).

In addition to elevated levels of vascular endothelial growth factor (VEGF) and fibroblast growth factor (FGF), stimulated peripheral whole blood from individuals with MDD was shown to have an increased PDGF production (Rachayon et al., 2022). Patients with MDD have elevated CSF GFAP levels (Michel et al., 2021) and serum GFAP levels that may rise with increasing MDD severity (Steinacker et al., 2021). Plasma GFAP levels are also elevated in individuals with neuropsychiatric illnesses who lack brain radiological abnormalities (Esnafoglu et al., 2017). Decreased GFAP expression has been observed in post-mortem cerebral tissues from patients with MDD (Michel et al., 2021; Rajkowska et al., 2018) and the expression of both GFAP mRNA and protein is decreased in astrocytes in the postmortem locus coeruleus of individuals with MDD (Chandley et al., 2013). GFAP mRNA is also decreased in the postmortem prefrontal cortex of depressed suicide victims (Nagy et al., 2015).

In addition, we discovered that the four neuroaxis biomarkers measured here were strongly correlated with the physio-affective phenome of MDD and that they had additive effects in predicting the severity of the physio-affective phenome. The latter is comprised of intertwined elevated scores on depression, anxiety, chronic fatigue, and physio-somatic (previously known as psychosomatic) symptoms, indicating that a common core, namely the overall severity of the physio-affective phenome of MDD, underlies these various manifestations of the phenome.

### Astroglial and neuronal projection toxicity in MDD

Most importantly, annotation and enrichment analysis showed that the enlarged networks of the four upregulated neuroaxis proteins were enriched in glial cell and neuronal projections, axons, the cytoskeleton, and axonal transport, including that of a mitochondrion. All-in all, it appears that MDD is characterized by increased serum biomarkers of disorders in astroglial and neuronal projections, including structural and functional aberrations in the axons, phenomena that we would propose to call astroglial & neuronal projection toxicity.

First, endothelial cells and perivascular mural cells express PDGFR and PDGR-BB (composed of two B subunits) in the brain (Vanlandewijck et al., 2018). PDGFRβ and its ligand PDGF-BB are involved in blood vessel formation and maintaining the integrity of the BBB (Kimura et al., 2020; Montagne et al., 2015). In AD, cleavage of PDGFR’s extracellular domain into the CSF disrupts PDGFR signaling, leading to BBB permeability, hypoperfusion, and cognitive impairment (Montagne et al., 2020). Because there is a strong link between CFS and serum levels (Miners et al., 2019), it is likely that peripheral serum levels of soluble PDGFR reflect brain levels.

Second, serum GFAP has been advocated as a marker to monitor astroglial pathology, including during the course of MDD (Al Shweiki et al., 2017; Hol and Pekny, 2015; Steinacker et al., 2021). CSF levels of GFAP may reflect astrocytic changes, and in neurodegenerative diseases, serum GFAP has been suggested to constitute a diagnostic or monitoring biomarker (Abu-Rumeileh et al., 2019; Oeckl et al., 2019). Alterations in GFAP levels may occur as a result of traumatic injuries and may appear during the course of neurodegenerative or neuroinflammatory processes (Abdelhak et al., 2019). Moreover, GFAP is involved in both long term depression and motor memory (Shibuki et al., 1996).

Third, NF-L levels are elevated in many neurological disorders and correlate with disease progression and brain atrophy in, for example, Alzheimer’s disease, multiple sclerosis, frontotemporal dementia, and amyotrophic lateral sclerosis (Lleó et al., 2019). Imaging studies have linked NF-L levels to changes in white matter fibers, general brain volume, and local atrophy, including in the hippocampus (Jakimovski et al., 2019; Khalil et al., 2020). In this respect, increased NF-L levels appear to represent ongoing and acute disease processes rather than cumulative brain damage in patients with neuroinflammatory illnesses (Cantó et al., 2019; Srpova et al., 2021). Furthermore, increased NF-L in neurodegenerative disoders is a biomarker of the cyto-axonal cell structure, which is a key component of the axon cytoskeleton (Gaetani et al., 2019; Khalil et al., 2018), and maybe synaptic degeneration (Zerr et al., 2018). The significant link between serum NF-L and cognitive performance in healthy people indicates that NF-L can identify microstructural changes at a subclinical level (Beste et al., 2019). Consequently, increased serum NF-L levels in MDD might be considered as a sign of active neuropathological processes (Dohm et al., 2017; Douillard-Guilloux et al., 2013) and more specifically of axonal and synapse injuries.

Fourth, serum P-tau217 (Janelidze et al., 2020; Telser et al., 2022) and CSF P-tau217 (Janelidze et al., 2020) have the potential to predict the progression of structural brain abnormalities, and both CSF and serum levels start to increase simultaneously (Palmqvist et al., 2019). Since tau stabilizes axonal transport and microtubule dynamics and maintains synaptic integrity (Tracy et al., 2022; Wang et al., 2022), it may be concluded that elevated P-tau217 levels indicate neuroaxis, axonal and synaptic injuries (Gaetani et al., 2019). Interstingly, there were significant associations between the NF-L, P-tau217 and GFAP and we were able to extract a principal component from the three biomarkers.

### Contributions of IR and calcium to the physio-affective phenome of MDD

The third most important finding of this research is that elevated IR and decreased calcium (but not magnesium) are observed in MDD and, together with the astroglial & neuronal projection-toxicity biomarkers and CRP, contribute to the severity of the physio-affective phenome of MDD. It is assumed that peripheral assessments of IR (such as HOMA-IR) reflect brain insulin sensitivity (Kullmann et al., 2016). IR causes endothelial dysfunctions (Janus et al., 2016), which generate increased permeability of the blood-brain barrier (BBB), allowing excessive entrance of proinflammatory cytokines into the CNS, culminating in neuroinflammation, neuronal malfunctions and neurodegeneration (Kleinridders et al., 2014). Fewer dendritic spines, lower BDNF levels, impaired synaptic plasticity, and neurodegeneration are associated with brain IR (Milstein and Ferris, 2021; Park et al., 2019). IR is related to decreased memory and executive functioning, as well as a decrease in metabolic activity in the medial prefrontal cortex, a reduction in hippocampal volume, and an aberrant intrinsic connection between the hippocampus and the medial prefrontal cortex (Kenna et al., 2013; Rasgon et al., 2011; Rasgon et al., 2014).

As indicated in the introduction, there is evidence that abnormalities in calcium metabolism may be linked to neurodegenerative processes and illnesses (Bezprozvanny, 2010). Calcium signaling dysregulation is suggested as one of the early stages in the etiology of several neurodegenerative illnesses (Bezprozvanny, 2010). However, there is no evidence to suggest that low blood calcium levels in serum have a mechanistic effect on these brain networks. Decreased blood calcium is a well-established biomarker of peripheral inflammation, as seen in acute COVID-19 infection and type 2 diabetes mellitus (Al-Hakeim et al., 2022a; Al-Jassas et al., 2022). Because of this, decreased calcium may be a secondary biomarker of the IRS response.

### Peripheral inflammation and IR contribute to neuro-glial projection toxicity

The fourth major finding of this study is that the levels of CRP and IR together explained 28.9% of the variance in the composite neuroaxis index. These results suggest that peripheral markers of inflammation and IR in MDD may affect astroglial and neuronal projections.

Peripheral inflammation affects the brain via humoral and neural pathways, altering neuronal cell function, structural and functional connectivity, and, as a result, behaviors such as depression (Harrison et al., 2016; Kubera et al., 2011). Multiple neuro-immune (M1, Th1, and Th17) and neuro-oxidative pathways may produce neurotoxicity, interfering with neuronal activities such as neurogenesis, neuroplasticity, receptor expression, axonogenesis, and dendritic sprouting, etc., according to the current thinking (Maes et al., 2022a; Maes et al., 2009). In addition, inflammation impairs functional connectivity within the brain network that is essential for interoceptive signaling, i.e., the transfer of peripheral body signals to the brain, which has implications for the etiology of inflammation-linked depression. (Aruldass et al., 2021).

As discovered in this study, CRP and IR may further affect the astroglial and neuronal projections through distinct mechanisms. As a result, peripheral activation of immune-inflammatory pathways and increased IR, together with neuro-oxidative toxicity and decreased antioxidant, neuroprotective, and CIRS defenses, may influence the projections (Maes, 2008, 2022; Maes et al., 2021). Importantly, the findings of this research indicate that this astroglial and neuronal projection toxicity not only influences the affective (depression and anxiety), but also the chronic fatigue and physio-somatic symptoms of MDD. Both symptom domains are often associated with the same biomarkers in MDD (Anderson et al., 2014; Maes et al., 2012).

Our results expand the neuroimmunotoxicity hypothesis of MDD, as outlined in the Introduction (Maes et al., 2022a; Maes et al., 2009; Schiepers et al., 2005b), and suggest that astroglial and neuronal projections are especially affected.

## Conclusion

A large part of the ariance in the physio-affective phenome of MDD is explained by biomarkers of neuroaxis injury, IR, and decreased calcium. In addition, CRP and IR predict part of the variance in the neuroaxis index. Annotation and enrichment analysis indicate the contribution of astroglial and neuronal projection toxicity in the pathophysiology of MDD. Inflammation, IR and lowered calcium as well as astroglial and neuronal projections appear to be new drug targets to treat the depression, anxiety, chronic fatigue and physiosomatic symptoms of MDD.

## Data Availability

The dataset generated during and/or analyzed during the current study will be available from Michael Maes upon reasonable request and once the dataset has been fully exploited by the authors.

## Author’s contributions

All authors contributed equally to this paper.

## ETHICS APPROVAL AND CONSENT TO PARTICIPATE

The “institutional ethics board of the University of Kufa” approved the study (617/2020).

## HUMAN AND ANIMAL RIGHTS

The study was conducted according to Iraq and international ethics and privacy laws and was conducted ethically in accordance with the World Medical Association Declaration of Helsinki. Furthermore, our IRB follows the International Guideline for Human Research protection as required by the Declaration of Helsinki, The Belmont Report, CIOMS Guideline and International Conference on Harmonization in Good Clinical Practice (ICH-GCP).

## CONSENT FOR PUBLICATION

All participants gave written informed consent before participation in this study.

## FUNDING

There was no specific funding for this specific study.

## Conflict of interest

The authors have no conflict of interest with any commercial or other association connected with the submitted article.

## Acknowledgments

We thank the staff of Al-Sadr Teaching Hospital and Al-Amal Specialized Hospital for Communicable Diseases in Najaf governorate-Iraq for their help in the collection of samples. We also thank the high-skilled staff of the hospital’s internal labs, especially Mr. Murtadha Al-Hilo, for their help in the estimation of biomarkers levels.

## Notes

### Competing Interest Statement

The authors have declared no competing interest.

### Author Declarations

ETHICS APPROVAL AND CONSENT TO PARTICIPATE The institutional ethics board of the University of Kufa approved the study (617/2020). HUMAN AND ANIMAL RIGHTS The study was conducted according to Iraq and international ethics and privacy laws and was conducted ethically in accordance with the World Medical Association Declaration of Helsinki. Furthermore, our IRB follows the International Guideline for Human Research protection as required by the Declaration of Helsinki, The Belmont Report, CIOMS Guideline and International Conference on Harmonization in Good Clinical Practice (ICH-GCP). CONSENT FOR PUBLICATION All participants gave written informed consent before participation in this study.

